# Neurodevelopmental copy number variants increase risk of internalising and cardiometabolic multimorbidity: findings from UK Biobank

**DOI:** 10.1101/2025.05.13.25326577

**Authors:** Ioanna K Katzourou, LINC consortium, Inês Barroso, Julie Clayton, Golam Khandaker, Daniel Stow, Nicolas Timpson, Ruby Tsang, Jack Underwood, Megan Wood, George Kirov, James Walters, Michael J Owen, Peter Holmans, Marianne B M van den Bree

## Abstract

**Background:** Internalising and cardiometabolic multimorbidity (ICM-MM) represents a major clinical challenge, negatively impacting life expectancy and quality of life and resulting in considerable healthcare costs. Individuals with a copy number variant associated with increased risk of neurodevelopmental conditions (ND-CNV) are more likely to develop mental or physical ill health, however the effects on ICM-MM remain poorly understood.

**Methods:** We used data from UK Biobank (ND-CNV N= 7,549, 1.62%). ICM-MM was identified using electronic healthcare records. It was defined as combinations of any internalising condition (either depression, anxiety or somatoform disorder) with each of five cardiometabolic conditions (hypertension, dyslipidemia, obesity, type 2 diabetes and chronic kidney disease). We also studied whether ICM-MM risk in those with ND-CNV differed by sex, presence of a deletion versus a duplication and we established associations of dosage-sensitive genes within ND-CNVs with ICM-MM. Finally, we explored the interaction between presence of ND-CNV and polygenic risk scores (PRSs) of internalising and cardiometabolic traits on ICM-MM risk.

**Results:** The presence of ND-CNV was associated with ICM-MM (OR range 1.21-1.57). Female participants with ND-CNV were more likely to have any internalising condition and T2D, and those with a deletion any internalising condition and obesity. The number of deleted haploinsufficient genes, but not duplicated triplosensitive genes, was associated with ICM-MM. No significant interactions between ND-CNVs and PRSs were found.

**Conclusions:** This is the first study to report that ND-CNVs increase the likelihood of ICM-MM, and we find evidence of sex differences and stronger effects for deletions. Increased clinical awareness can help ameliorate this risk.

## Background

Multimorbidity, also referred to as multiple long-term conditions (MLTC), indicates the presence of two or more chronic health conditions in the same individual. Multimorbidity represents a major public health concern, with at least 50 million people affected in the European Union alone(1). An estimated 25% of the population of high-income countries is living with two or more conditions, and rates are rapidly increasing in low- and middle-income countries(1,2). Multimorbidity is associated with high personal as well as societal healthcare costs(2). Moreover, multimorbidity is difficult to manage, resulting in high risk of failure of care and putting considerable strain on healthcare systems(1–3).

Cardiometabolic conditions, such as hypertension, obesity and type 2 diabetes (T2D), and internalising conditions, such as depression and anxiety, are highly prevalent, affecting millions of people worldwide and resulting in increased disability and functional decline, poor quality of life and premature mortality(4,5). The co-occurrence of internalising and cardiometabolic conditions represents the most common type of physical and mental health multimorbidity in older adults(3). While both cardiometabolic and internalising conditions pose considerable public health challenges, their presence in combination is particularly burdensome for the individuals affected (4,5). Internalising disorders are associated with an increased risk of subsequent cardiometabolic disorders(6–8) as well as cardiovascular morbidity and premature mortality(9,10). The reverse is also the case, with those with cardiometabolic conditions experiencing more depression and anxiety(7,10–12). Importantly, the lower life expectancy associated with depression(13) is partially attributable to comorbid cardiovascular disease(14). These findings highlight the importance of uncovering the risk factors contributing to internalising and cardiometabolic multimorbidity (ICM-MM).

While the exact mechanisms leading to the development of ICM-MM are unknown, shared genetic risk factors may play a considerable role(15,16). Polygenic risk scores (PRS) of cardiometabolic disorders have been found to be associated with depression(17,18), while high genetic overlap has also been found between depression and T2D(19). Most published studies focus on common genetic variation, while the effect of rare genetic variants on multimorbidity remains mostly unexplored. Copy number variants (CNVs) are structural alterations in chromosomes involving the deletion or duplication of a section of varying length(20). A range of CNVs increase the risk of neurodevelopmental conditions (NDCs), such as intellectual disability, autism spectrum disorder, attention deficit hyperactivity disorder and schizophrenia(21–24). These are referred to as ND-CNVs(25). In addition to NDCs, these CNVs can also increase the risk of internalising conditions(26) as well as a range of physical health conditions and traits(27), including hypertension, diabetes and body mass index (BMI). There is increasing evidence of increased risk of cardiovascular morbidity in individuals with some of these variants(28–30). As ND-CNVs are often diagnosed in childhood, increased understanding of the development of multimorbidity in individuals with ND-CNVs presents opportunities for better understanding the development of disease development and potentially also provide important insights into the biological mechanisms of multimorbidity.

In this study, we explored the association between ICM-MM and ND-CNVs(21) in UK Biobank (UKBB)(31).

The objectives of this study were to:

- Determine the association of ND-CNVs, loss (deletion) and gain (duplication) of chromosomal material and dosage-sensitive genes in ND-CNVs with ICM-MM;
- Investigate sex diherences in the ehect of ND-CNVs on ICM-MM;
- Establish if the ehect of common variation (PRSs of internalising and cardiometabolic traits) on ICM-MM dihers in individuals with versus without a ND-CNV.

## Methods

### Data source

UKBB is a prospective cohort of over 500,000 individuals living in the United Kingdom(31). UKBB received ethical approval from the North West - Haydock Research Ethics Committee (reference 16/NW/0274). Participants provided electronic signed consent at recruitment. This study was conducted under application number 79704.

### Participants

Participants aged between 40 and 69 years old were recruited into UKBB between 2006 and 2010(31). Sociodemographic, lifestyle and medical history information were collected using touchscreen questionnaires. Physical and functional measurements, biochemical assays and genome-wide genotyping were collected at a baseline assessment. Linkage to the National Health Service provided data on deaths, cancer diagnoses, hospital inpatient/outpatient episodes and primary care records. Details of the UKBB study design are provided elsewhere(31). Out of the total number of UKBB participants (502,411), 459,483 (91.47%) had available CNV call data and were included in this study.

### Phenotyping

This study took place under the Lifespan Multimorbidity Research collaborative (LINC) (https://www.cardiff.ac.uk/lifespan-multimorbidity-research-collaborative). LINC seeks to understand the development of internalising and cardiometabolic multimorbidity (ICM-MM) over the life course. The conditions included in LINC’s definition of ICM-MM were selected following discussions with primary and secondary care doctors, the LINC patient and public involvement team, as well as the LINC team of researchers and clinicians.

Three internalising conditions were included: depression, anxiety and somatoform disorder. Five cardiometabolic conditions were included: hypertension, dyslipidemia, obesity, T2D and chronic kidney disease (CKD). The cardiometabolic conditions included in the definition are restricted to those that generally occur earlier in life than established cardiovascular disease, allowing the opportunity to study the increase in risk over an extended period.

Individuals with each of the conditions of interest were identified through linked electronic healthcare records (EHR; primary care records and hospital episode statistics (HES)) using established clinical codelists (see Supplementary Material). We examined the pairwise combinations of aggregated internalising conditions, defined as the presence of one or more of the three internalising conditions listed above (referred to as ‘any internalising condition’ from here onwards), and each of the five cardiometabolic conditions. We also constructed an outcome variable - any ICM-MM - defined as the presence of one or more of the three internalising conditions and one or more of the five cardiometabolic conditions.

### CNV calling

The process of the calling of the NDD-CNVs in UKBB is described in detail elsewhere(32). Briefly, calling was performed using PennCNV-Affy 1.0.3 protocols(33) on the UKBB genotype array data. Samples were excluded if they carried 30 or more CNVs, had a waviness factor greater than 0.03 or less than –0.03; a single-nucleotide polymorphism call rate lower than 96%; or log R ratio SD higher than 0.35. CNVs were excluded if they were covered by fewer than 20 probes; had a density coverage of less than 1 probe per 20,000 base pairs; or a confidence score lower than 10. This resulted in 459,483 individuals with available CNV call data.

### Statistical analyses

All statistical analyses were performed in R(34).

### Aggregated CNVs

We focused on a set of 54 CNVs that show strong evidence of increasing the risk of developing a neurodevelopmental disorder (ND-CNVs)(21). We assessed the association of the presence of any ND-CNV (aggerated ND-CNV; binary variable) with each of the conditions of interest individually as well as pairwise combinations of any internalising conditions and each cardiometabolic condition and any ICM-MM (binary variables). We used logistic regression of each of these phenotypes on aggregated ND-CNV, adjusting for age at baseline, sex and Townsend deprivation index (as a measure of socioeconomic status) and the first five genetic principal components to account for population stratification.

### Sensitivity analyses

While HES data are available for the whole UKBB cohort (N = 502,390), primary care records are available for ∼40% of the participants (N = 229,951). A sensitivity analysis was performed including only participants for whom both HES and primary care records were available (N= 229,951).

Deletions in the 16p11.2 region have been previously associated with class III obesity (formerly known as morbid obesity)(35–37). Obesity is one of the five cardiometabolic conditions we examined, as well as a known risk factor for the remaining four. In order to determine if the overall effect of ND-CNV on ICM-MM was driven by these specific ND-CNVs, we repeated the regression analysis after removing individuals with a proximal or distal deletion of 16p11.2 (N= 185). We also repeated the regressions adjusting for BMI at baseline (excluding obesity as an outcome) to account for possible effects of the ND-CNVs on body mass.

### Post-hoc analyses

To assess if there are any sex differences in the associations described above, we also conducted the above regression including an interaction term between ND-CNV and sex. Moreover, we also assessed the association of the phenotypes with the type of NDD-CNV, e.g. loss (deletion) or gain (duplication). For this analysis, genotype was coded as a three-level variable (no ND-CNV, deletion or duplication). Logistic regressions of each of the phenotypes on genotype were performed, adjusting for age, sex Townsend deprivation index and the first five genetic principal components. Individuals with both a deletion and a duplication were excluded from this analysis (N= 12).

We next examined if dosage-sensitive genes within ND-CNVs are associated with ICM-MM. The coordinates for each ND-CNV were used to map which genes were affected in each individual. Out of the genes affected by a ND-CNV in each individual, we identified those that are haploinsufficient (deletion intolerant) or triplosensitive (duplication intolerant), based on a dosage sensitivity analysis conducted by Collins *et al.*(38) and calculated the number of dosage sensitive genes affected in each individual. For this analysis, deletions and duplications were analysed separately. Thus, we regressed the phenotypes of interest (e.g. pairwise combinations of any internalising conditions with each cardiometabolic condition and any ICM-MM) on the number of haploinsufficient genes included in each deletion or the number of triplosensitive genes included in each duplication, adjusting for age, sex, Townsend deprivation index, the total number of genes included in each ND-CNV as a proxy of overall biological effect of each ND-CNV, and the first five genetic principal components to account for population stratification.

### Interaction between CNVs and common genetic variation

To assess whether the association of common genetic variation and the risk of multimorbidity differs in individuals with and without a ND-CNV, the evidence for interaction between the presence of a ND-CNV and PRSs of our internalising and cardiometabolic conditions of interest was assessed. PRSs for major depressive disorder (MDD)(39), anxiety(40), low density cholesterol (LDL; as a proxy for dyslipidemia)(41), body mass index (BMI; as a proxy of obesity)(42), systolic blood pressure (SBP; as a proxy for hypertension)(43), T2D(44) and CKD(45) were computed. The genome-wide association studies (GWAS) used for the generation of these PRSs were selected because they were the latest and largest studies with publicly available summary statistics that excluded UKBB. They are described in Supplementary Table 1.

PRS-CS(46) was used for PRS calculation. PRS-CS is a Bayesian algorithm that can infer posterior effect sizes of SNPs via continuous shrinkage(46), therefore avoiding the need for linkage disequilibrium pruning and p-value thresholding. The inferred posterior effect sizes were used for PRS generation on PLINK 2.0(47). In order to produce PRSs that are on the same scale across individuals from different ancestries, we adjusted them for ancestral differences in mean and variance using the 1000Genomes dataset as reference, as described by Khan et al.(48). The ancestry adjustment is described in detail elsewhere(49).

Logistic regression analyses were performed as described previously, including the main effects of each ancestry-adjusted PRS, ND-CNV and the interaction term PRS*ND-CNV.

### Individual CNVs

We assessed the association of the combination of aggregated internalising conditions with each cardiometabolic condition and also any ICM-MM with each of the ND-CNVs individually. The number of individuals with each of the ND-CNVs are shown in Supplementary Table 2. To ensure the analysis was statistically viable, CNVs that were observed fewer than 5 times in the total sample were excluded, resulting in 33 CNVs included in this analysis. The associations were assessed using logistic regression, adjusting for age at baseline, sex, Townsend deprivation index and the first five genetic principal components.

## Results

### Associations of aggregated ND-CNVs with ICM-MM

There were 7,546 individuals (1.64%) with a ND-CNV in UKBB. The demographic characteristics of the individuals with and without a ND-CNV are shown in Supplementary Table 3. Individuals with a higher Townsend deprivation index were more likely to have a ND-CNV (OR = 1.55, p-value = 1.19×10^-34^), meaning individuals with a ND-CNV were more likely to live in a more deprived environment than those without. The number of individuals with and without a ND-CNV that have a diagnosis of the eight conditions of interest is shown in Supplementary Table 4.

14.2% of individuals with a ND-CNV had ICM-MM, compared to 11.5% of individuals without a ND-CNV. The number of individuals with and without a ND-CNV that have ICM-MM is shown in Table 1.

**Table 1.**
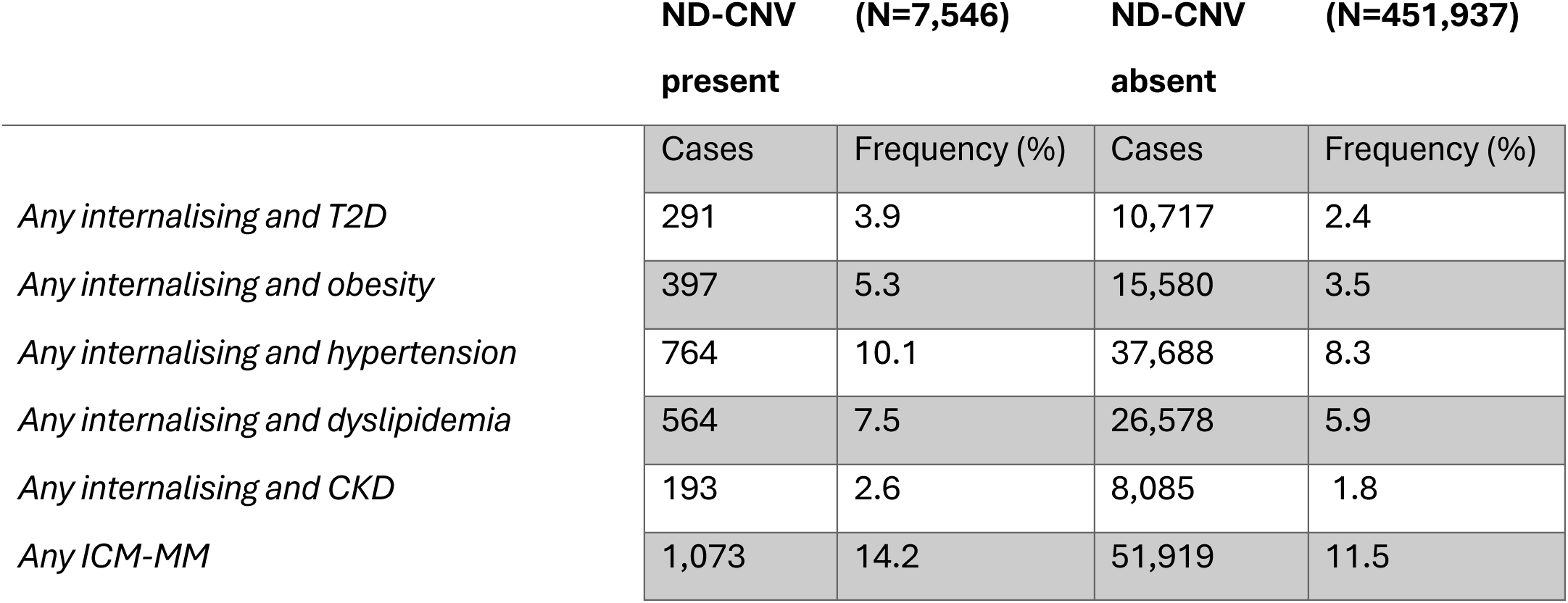
Counts and frequency for each of the ICM-MM phenotypes of interest in individuals with and without a ND-CNV.

All combinations of ICM-MM were significantly associated with the presence of a ND-CNV after Bonferroni correction for multiple testing (Bonferroni p-value threshold = 8.33×10^-3^). The OR for any ICM-MM was similar to that for any internalising condition or any cardiometabolic condition, however the ORs for certain ICM-MM combinations (any internalising conditions and obesity, any internalising condition and T2D, any internalising condition and CKD) were increased over any internalising condition or any cardiometabolic condition separately. The associations between the presence of a ND-CNV and the different combinations of aggregated internalising conditions with each cardiometabolic condition and ICM-MM are shown in Figure 1 and Supplementary Table 5.

**Figure 1.**
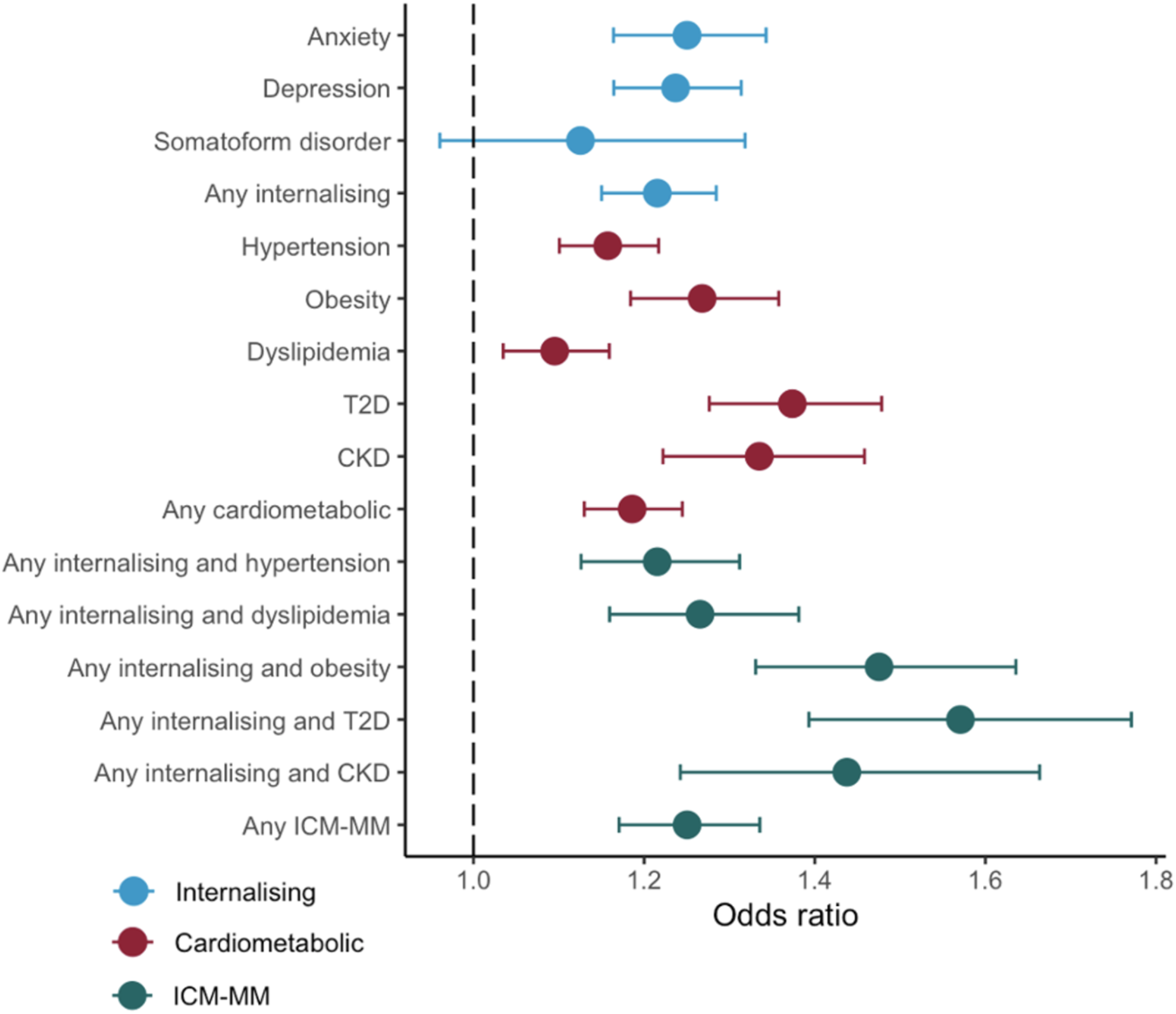
Association of ND-CNVs with individual conditions and multimorbidity.

The associations remained significant when adjusting for BMI (Supplementary Table 6). Restricting the analysis to individuals with both primary care and HES data (N= 229,951) did not substantively alter our findings (Supplementary Figure 1 and Supplementary Table 7), while the ORs were higher and p-values lower when including the full UKBB sample. Excluding individuals with proximal and distal deletions in 16p11.2 (N = 185), a known risk factor for early onset morbid obesity, also did not substantively alter the findings (Supplementary Figure 2 and Supplementary Table 8).

We examined whether the associations between having any ND-CNV and the phenotypic presentations of interest differed by sex. There was evidence of interaction between aggregated ND-CNV and sex in association with any cardiometabolic condition and the combination of any internalising condition and T2D (Figure 2 and Supplementary Table 9). Supplementary Figure 3 shows that the increased risk of T2D or hypertension associated with a ND-CNV is higher in female than in male participants.

**Figure 2.**
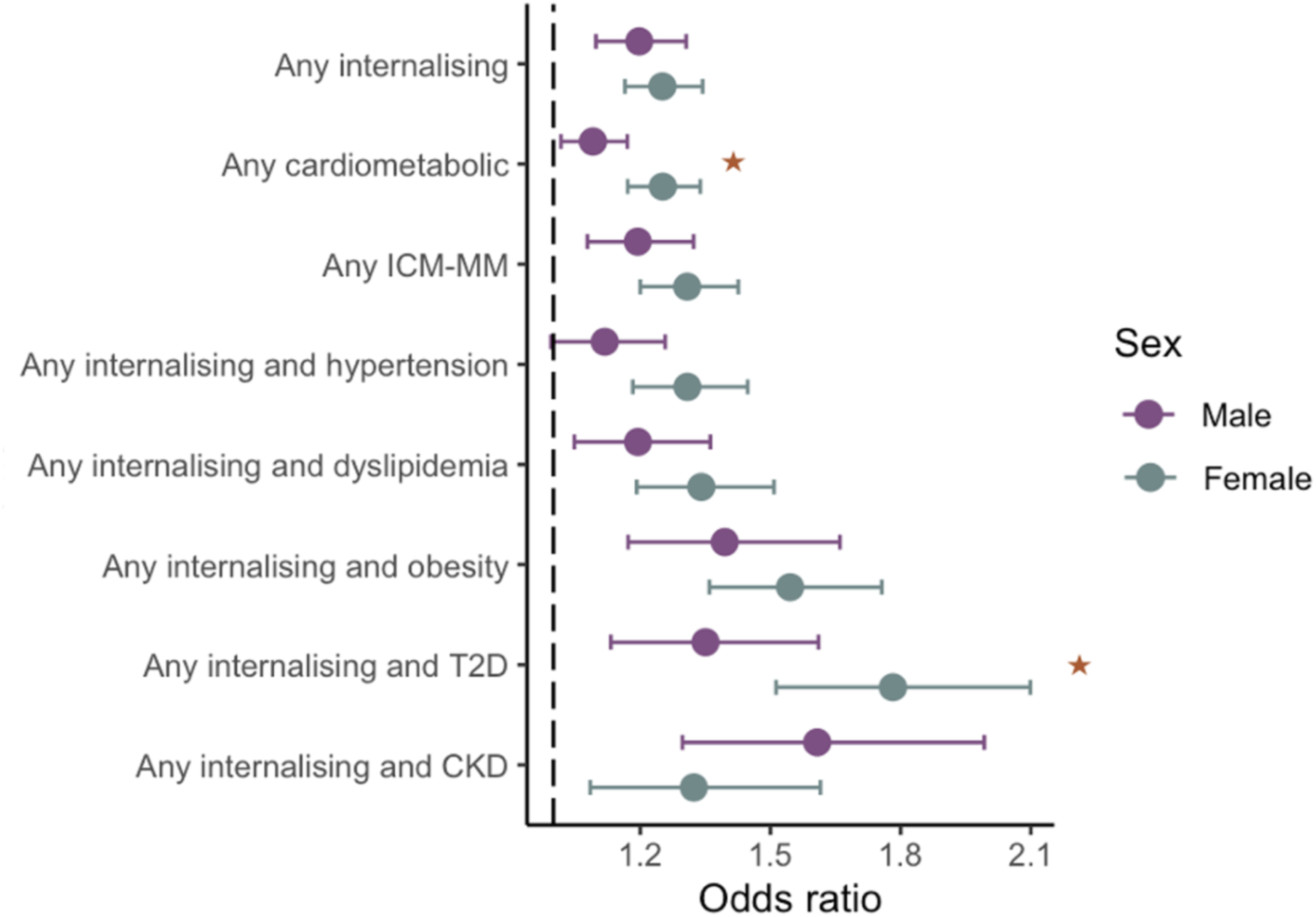
Association of ND-CNVs with multimorbidity for male and female sex. Stars indicates a significant interaction between the presence of ND-CNV and sex (p-value < 0.05).

We examined whether the associations between the outcomes of interest differed for individuals with loss (deletion) or gain (duplication) of chromosomal material. Having a deletion increased the risk of having internalising condition and obesity over having a duplication (Figure 3 and Supplementary Table 10). When considering individual conditions, having a deletion was associated with greater the odds of having obesity and T2D over having a duplication while having a duplication significantly was associated with greater odds of having dyslipidemia over having a duplication, as seen in Supplementary Figure 4.

**Figure 3.**
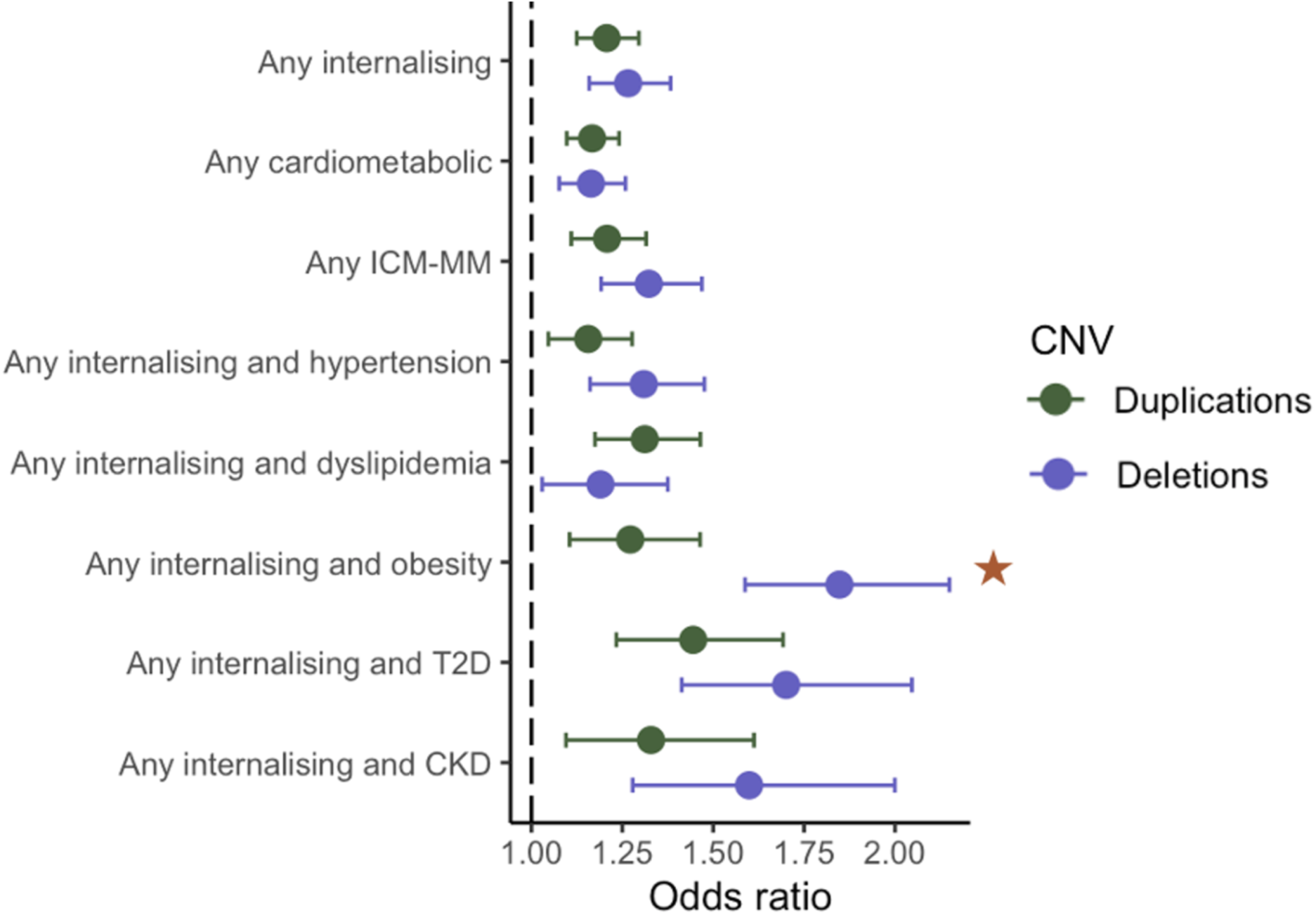
Association of duplications and deletions with multimorbidity. Stars indicate a suggested di;erence between deletions and duplications (p-value < 0.05).

The number of haploinsufficient genes within a deletion was significantly associated with all outcomes. The number of triplosensitive genes within a duplication did not show the same pattern (Supplementary Figure 5). The total number of genes affected by a deletion or a duplication was significantly associated with all outcomes (Supplementary Table 11).

### Interaction between aggerated ND-CNVs and common genetic variation

The evidence for association between ND-CNV and combinations of ICM-MM remained significant when adjusting for each PRS (and vice versa). However, interactions between PRS and aggregated ND-CNV were not significant after Bonferroni correction for multiple testing, with three interactions reaching nominal significance (p-value < 0.05; any internalising condition and obesity and any internalising condition and CKD for anxiety PRS, and any internalising condition and CKD for LDL PRS), as seen in Supplementary Table 12. PRSs of MDD (OR = 1.001, p-value = 1.34×10^-4^), anxiety (OR = 1.001, p-value = 2.40×10^-6^) and CKD (OR = 1.001, p-value = 2.54×10^-3^) were associated with the presence of a ND-CNV.

### Associations of individual ND-CNVs with internalising and cardiometabolic multimorbidity

For each of the 33 ND-CNVs that were present more than 5 times (Supplementary Table 1) in individuals in UKBB, we investigated their association with the pairwise combinations of any internalising condition with each cardiometabolic condition, as well as any ICM-MM (Supplementary Figure 6 and Supplementary Table 13). After Bonferroni correction for multiple testing, 9 of the 33 NDD-CNVs showed evidence for association with ICM-MM (p-value < 2.52×10^-4^). Several deletions on the p arm of chromosome 16 were associated with multiple ICM-MM outcomes, while 22q11.2 duplication, 15q24 duplication and 15q13.3 deletion were associated with any internalising condition and obesity. All significant associations found indicated an increased risk of ICM-MM.

## Discussion

The impact of ND-CNVs on risk of multimorbidity between mental and physical health conditions has received little research attention to date. The aim of this study was to investigate the association of ND-CNVs with the presence of ICM-MM in a population cohort. Our findings indicate that individuals with ND-CNVs are more likely to experience ICM-MM, with particularly increased odds for specific combinations of internalising and cardiometabolic conditions such as any internalising condition and T2D, any internalising condition and CKD and any internalising condition and obesity. We also found evidence of differential associations with ICM-MM in individuals with ND-CNVs depending on sex and whether they carry a deletion or duplication, and that genes intolerant to deletion (haploinsufficient genes) within ND-CNVs were more likely to contribute to ICM-MM risk than genes intolerant to duplication (triplosensitive genes). Finally, we found no evidence of interactions between ND-CNV status and PRSs of the internalising and cardiometabolic conditions under study, indicating that common genetic variation does not modify risk of ICM-MM in individuals with a ND-CNV.

We found that having any ND-CNV was associated with 25% increased likelihood of ICM-MM, with the frequency of any ICM-MM in individuals with a ND-CNV being 14.2%, compared to 11.5% in individuals without a ND-CNV. Given the way UKBB participants were ascertained(50), this is likely to be a lower bound estimate, as both the outcomes of interest and ND-CNVs are associated with premature mortality and the older age of participants could introduce a survival bias. While ND-CNVs have mostly been studied in relation to NDCs(51–53) and other psychiatric conditions(25,26,54), they have also recently been linked to multiple physical health outcomes(55,56), including obesity(35,57), renal disease(58) and diabetes(59). Whilst much of the literature linking ND-CNVs with physical health outcomes has focused on clinical populations and rare and severe syndromic phenotypes, we studied a population cohort that is not enriched for more severe manifestations of ND-CNVs. Whilst unknown, it is also likely that many of the individuals with ND-CNVs in UKBB are unaware of their genotype. We explored common, earlier onset cardiometabolic conditions that are on the causal pathway to cardiovascular morbidity and mortality, and we found that ND-CNVs were significantly associated with all the cardiometabolic conditions we examined, as well as ICM-MM. Our results suggest that ND-CNVs are associated with common conditions and ICM-MM even in the general population, while it is important to keep in mind that these outcomes are likely to be more severe in clinically ascertained carriers. Associations between ND-CNVs and ICM-MM remained significant when removing ND-CNVs that are known risk factors for morbid obesity or when correcting for BMI, suggesting that in this dataset the associations are unlikely to be driven by the effect of ND-CNVs on body mass.

In addition to our composite phenotype of any ICM-MM, we also assessed the impact of ND-CNVs on pairwise combinations of any internalising condition and each of the five cardiometabolic conditions. Our rationale was that while the internalising disorders are highly phenotypically and genetically correlated(49,60,61), the cardiometabolic disorders are more heterogeneous, therefore the associations with ND-CNVs may differ between the pairwise ICM-MM combinations. Our findings confirm this assumption. While the odds for any ICM-MM conferred by ND-CNVs was quite similar to those for any internalising condition or any cardiometabolic condition alone, the odds for some of the combinations (e.g. any internalising condition and obesity, any internalising condition and T2D, any internalising condition and CKD) was higher than those for any internalising condition, any cardiometabolic condition or any ICM-MM. This indicates that individuals with ND-CNVs have higher odds of developing certain combinations of internalising and cardiometabolic conditions than others and could benefit from closer monitoring for early indications of particular conditions.

Interestingly, we observed that the effect of the presence of a ND-CNV on the odds of any cardiometabolic condition as well as any internalising condition and T2D appeared to be higher in female than in male participants. While there was no strong evidence for the remaining outcomes, there was a pattern for larger effects for female than for male participants for all ICM-MM combinations apart from any internalising condition and CKD. Previous studies on sex differences in the effect of ND-CNVs on NDCs and other psychiatric disorders have produced conflicting results(26,62–65), with some studies reporting higher rates of depression in female adults with ND-CNVs(26) and of anxiety and depression in female children with with a diagnosed NDC and an ND-CNV(65), whilst others found no evidence of sex differences in ND-CNV burden for anxiety and depression(62). While further studies are required to understand the underlying mechanisms behind possible sex differences, our findings highlight the importance of considering sex as a factor when designing tailored health monitoring strategies for individuals with ND-CNVs.

Moreover, we found suggestive evidence of differences between the effect of deletions and duplications on ICM-MM, with deletions being associated with higher odds of having any internalising condition and obesity over duplications. Deletions in the 16p11.2 region are known risk factors for class III obesity(35–37), and it is likely that these regions contribute to this finding, however the association between ND-CNVs and any internalising condition and obesity remained significant when deletions in this region were removed, suggesting that other ND-CNVs also predispose to this type of multimorbidity. In order to further explore the way in which ND-CNVs are involved in the pathogenesis of multimorbidity, we examined the association of dosage-sensitive genes within a ND-CNV with ICM-MM. Haploinsufficient genes (those intolerant to deletion) within ND-CNVs were associated with higher odds of most combinations of ICM-MM, whereas triplosensitive genes (those intolerant to duplication) did not show evidence of strong effect. The total number of genes within both deletions and duplications was significantly associated with most combinations of ICM-MM, however the effect size was small. Our findings suggest that, for the phenotypes we studied, in individuals with deletions the loss of haploinsufficient genes is pathogenic, however the same might not be the case for duplications, where the total number of genes hit or other parameters, like the total length or the penetrance of the variant, might be more important than the number of triplosensitive genes duplicated. Importantly, for individuals with deletions, the number of haploinsufficient genes can be used to identify individuals that might be at risk of developing a more severe phenotype.

We explored the possibility of interaction between the presence of a ND-CNV and PRSs of our internalising and cardiometabolic traits of interest on risk of ICM-MM. The absence of detectable interaction between ND-CNVs and the PRSs suggest that effect of ND-CNVs on the risk of ICM-MM is not modified by the effect of common genetic variation. This is the first study to explore the interaction of common variation with ND-CNVs on the risk of multimorbidity. Although UKBB is one of the largest cohorts internationally available for the study of our aims, ND-CNVs are rare (they were present in 1.6% of participants) and it is likely that we lacked sufficient statistical power to detect possible interactions, however our results are in agreement with previous studies exploring the effect of CNVs or other rare variants and common variation on depression phenotypes in cohorts of similar or smaller size (66–68).

When examining the ND-CNVs individually, we found nine with evidence for association with at least one type of ICM-MM. Four of these were deletions on the p arm of chromosome 16 (16p13.11 deletion, 16p11.2 deletion, 16p12.1 deletion and 16p12.1 distal deletion). 16p11.2 deletion, 16p12.1 deletion and 16p13.11 deletion were associated with all combinations of ICM-MM, while 16p12.1 distal deletion was associated with any internalising condition and T2D and any internalising condition and obesity. These results are consistent with previous research finding that deletions in this region are associated with psychiatric disorders and traits(25,51), obesity(35–37), T2D(36) and kidney disease biomarkers(36). 22q11.2 duplication was associated with any internalising condition and obesity. This variant is considered to be associated with a variable and mild phenotype compared to deletions in the same region(69). Our results highlight the considerable heterogeneity between ND-CNVs. Where increasing sizes of population-based cohorts allow the opportunity, each CNV should be studied in isolation to better understand its specific contribution to disease risk and provide valuable insight into the biological underpinning of ICM-MM.

Our findings give evidence of association between ND-CNVs and the co-occurrence of common internalising and cardiometabolic conditions; a multimorbid presentation that can lead to substantive functional impairment, a reduced quality of life and premature mortality. This finding has several clinical implications. Firstly, it underscores the need for healthcare systems to adopt a more holistic approach to multimorbidity, particularly in relation to individuals with rare genetic variants. Current healthcare service provision tends to look at each diagnosis in isolation, but it is evident from our findings that this approach may not be suited for individuals with ND-CNVs, who are at risk from multiple conditions throughout their lifetime, many of which need to be managed by different medical specialties. It is imperative that healthcare systems transition to a more comprehensive multidisciplinary model of care, particularly for individuals with known pathogenic genetic variants. Secondly, our results suggest that the impacts of ND-CNVs extend beyond the phenotypes they are traditionally associated with (e.g. congenital abnormalities, intellectual disability, autism, schizophrenia) to include common morbidities and multimorbidity seen in older age people in the general population. Recent breakthroughs in sequencing technology allow for identification of an increasing range of rare genetic variants by medical genomics services, and children with NDCs will often be screened for ND-CNVs. ND-CNVs are often inherited and reduced penetrance may mean that seemingly unaffected relatives might carry an ND-CNV that predisposes them to ICM-MM, and could also stand to benefit from screening.

There are some limitations to our study. UKBB is one of the largest publicly available population cohorts, and was designed as a prospective study of middle and older age(31), with median age at recruitment being 57 years. It is therefore well-suited to this study as the age range of the participants allowed us to study conditions that tend to appear later in life. However, it is also susceptible to survival bias. As multimorbidity and CNV burden are associated with premature mortality(56,70), it is likely that individuals with the most severe outcomes are underrepresented, which would lead to effective reduction multimorbidity frequency and effect sizes compared to the general population. Moreover, the participants of UKBB have been found to have higher socioeconomic status, report higher overall health and be less ethnically diverse than the average UK population(71), which could also lead to reduced population-based effect sizes. Furthermore, the generalisability of our findings remains uncertain, particularly given the predominance of individuals of European descent in the UKBB. When assessing the interaction between ND-CNVs and PRSs, we found that some of the PRSs were associated with ND-CNV status (MDD, anxiety and CKD PRS). These associations could introduce a collider bias in the analysis, whereby adjusting for PRS would alter the ND-CNV association with the outcomes in a manner that resembles an interaction. However, we did not find evidence of interaction between ND-CNVs and any of the PRSs in our analysis. Finally, the variants we examined tend to be low in frequency, some appearing less than 100 instances in half a million of UKBB participants. We aggregated ND-CNVs to have increased statistical power, which facilitated discovering associations that would otherwise be missed. This approach provides valuable insights into the common effects of the variants but does not allow clarification of ND-CNV-specific relationships.

In conclusion, this study demonstrates that, on a population level, individuals with ND-CNVs have higher odds of developing common and preventable forms of ICM-MM. As the field of medical genetics continues to expand, understanding how rare genetic variants contribute to multimorbidity will be essential for improving the health outcomes of affected individuals.

## Acknowledgements

This research has been conducted using the UK Biobank Resource under Application Number 79704.

This work was funded by the Tackling Multimorbidity at Scale Strategic Priorities Fund programme (MR/W014416/1) delivered by the Medical Research Council and the National Institute for Health Research in partnership with the Economic and Social Research Council and in collaboration with the Engineering and Physical Sciences Research Council. IKK is supported by this grant.

Full list of LINC members:

Marianne B. M. van den Bree, George Kirov, Michael J. Owen, James T. R. Walters, Peter A. Holmans, Jane Lynch, Ioanna K. Katzourou, Jack F. J. Underwood (Cardiff University, UK)

David A. van Heel, Sarah Finer, Daniel Stow (Queen Mary University of London, UK)

Golam M. Khandaker, Nicholas J. Timpson, John A. A. MacLeod, Julie P. Clayton, Ruby S. M. Tsang, Jane Sprackman, Shahid Khan (University of Bristol, UK)

Inês Barroso, Rupert A. Payne (University of Exeter, UK)

Mark Mon-Williams, Megan L. Wood, Nabila Ali (University of Leeds, UK)

Hilary C. Martin (Wellcome Sanger Institute, UK)

Thomas Werge, Andrés Ingason, Morteza Vaez, Lam O. Huang (Institute of Biological Psychiatry, Denmark)

We thank the members of the LINC study public advisory group for their contribution.

## Funding

This work was funded by the Tackling Multimorbidity at Scale Strategic Priorities Fund programme (MR/W014416/1) delivered by the Medical Research Council and the National Institute for Health Research in partnership with the Economic and Social Research Council and in collaboration with the Engineering and Physical Sciences Research Council.

## Author contribution

Study conceptualisation and design: IKK, MvdB, PH, GK, MJO, JW, LINC. Analytical consultation and interpretation: IKK, MvdB, PH, GK, MJO, JW, AI, RT, DS, IB. UKBB data curation: IKK. Genetic data preparation: IKK. Supervision: MvdB, PH, GK, MJO, PH, JW. All listed authors critically edited the manuscript.

## Data availability

Relevant data is available from UK Biobank subject to standard procedures (www.ukbiobank.ac.uk).

## Declaration of interest

Michael J Owen is receiving research grants from Takeda Pharmaceuticals and Akrivia Health. The remaining authors declare no competing interests.

