## Supplementary Material for "Neurodevelopmental copy number variants increase risk of internalising and cardiometabolic multimorbidity: findings from UK Biobank"

The lists of clinical codes curated by the MULTIPLY project were used to establish instances of the conditions of interest(1). The primary care codelists for depression and anxiety were amended by psychiatrists within the LINC team. Dyslipidemia is not defined within the MULTIPLY clinical code lists, therefore the clinical code list from Baksh *et al.* was used to identify instances of dyslipidemia within primary care records(2), and ICD-10 codes E78.0, E78.1, E78.2, E78.3, E78.4, E78.5, E88.81 were used to define dyslipidemia in HES.

| Trait | GWAS |
|---|---|
| MDD | Wray *et al.,* 2018 (UKBB removed) (3) |
| Anxiety | Meier *et al.,* 2019 (4) |
| LDL | Willer *et al.,* 2013 (5) |
| SBP | Keaton *et al.,* 2024 (6) (only ICBP) |
| BMI | Locke *et al*. 2015 (7) |
| T2D | Mahajan *et al.,* 2018 (UKBB removed) (8) |
| CKD | Pattaro *et al.,* 2016 (9) |

*Supplementary Table 1. Genome-wide association studies used for polygenic risk score generation.*

| CNV | Count |
|---|---|
| Any | 7,549 |
| *1p36 del* | 1 |
| TAR del | 80 |
| TAR dup | 463 |
| 1q21.1 del | 119 |
| 1q21.1 dup | 193 |
| NRXN1 del | 176 |
| 2q11.2 del | 34 |
| 2q13 del | 56 |
| 2q13 dup | 73 |
| *2q37 del* | 1 |
| 3q29 del | 9 |
| *WH dup* | 3 |
| *WBS del* | 1 |

| | |
|---|---|
| WBS dup | 16 |
| *8p23.1 del* | 4 |
| 8p23.1 dup | 8 |
| *EMHT1 dup* | 1 |
| *10q23 del* | 3 |
| 15q11.2 del | 1,748 |
| 15q11.2 dup | 2,284 |
| *PWS del* | 1 |
| PWS dup | 19 |
| 15q13.3 del | 47 |
| *15q24 del* | 1 |
| 15q24 dup | 9 |
| *15q25 del* | 1 |
| 16p13.11 del | 140 |
| 16p13.11 dup | 888 |
| 16p12.1 del | 260 |
| 16p11.2distal del | 62 |
| 16p11.2distal dup | 142 |
| 16p11.2 del | 123 |
| 16p11.2 dup | 142 |
| 17p13.3 YWHAE del | 27 |
| 17p13.3 YWHAE dup | 8 |
| *17p13.3 PAFAH1B1 del* | 1 |
| *17p13.3 PAFAH1B1 dup* | 3 |
| *SMS* | 2 |
| Potocki Lupski | 6 |
| 17q11.2 delNF1 | 10 |
| 17q11.2 dup NF1 | 3 |
| 17q12 del | 9 |
| 17q12 dup | 104 |
| 22q11.2 del | 10 |
| 22q11.2 dup | 294 |
| 22q11.2 distal del | 5 |
| 22q11.2 distal dup | 14 |

*Supplementary Table 2. Number of individuals with each ND-CNV. ND-CNVs that appeared less than five times are in italics.*

| | ND-CNV | No ND-CNV |
|---|---|---|
| Age at recruitment (Mean (min, max)) | 56.07 (40,70) | 56.55 (37,73) |
| Townsend deprivation index (Mean (min, max)) | -0.86 (-6.26, 10.56) | -1.32 (-6.26, 11.00) |
| Female (N (%)) | 3,934 (52.11) | 245,589 (54.32) |

*Supplementary Table 3. Demographic characteristics of individuals with and without a ND-CNV. NV. Townsend deprivation index is measure of socioeconomic deprivation, with higher value indicating a more deprived environment.*

| | ND-CNV | (N=7,546) | No ND-CNV | (N=451,937) |
|---|---|---|---|---|
| | Cases | Frequency (%) | Cases | Frequency (%) |
| Anxiety | 878 | 11.6 | 42,365 | 9.4 |
| Depression | 1,335 | 17.7 | 65,040 | 14.4 |
| Somatoform disorder | 161 | 2.1 | 8,513 | 1.9 |
| Any internalising disorder | 1,685 | 22.3 | 84,755 | 18.8 |
| Hypertension | 2,765 | 36.6 | 153,829 | 34.0 |
| Obesity | 975 | 12.9 | 46,176 | 10.2 |
| Chronic kidney disease | 570 | 7.6 | 26,924 | 6.0 |
| Type II diabetes | 870 | 11.5 | 39,236 | 8.7 |
| Dyslipidemia | 1,709 | 22.6 | 95,960 | 21.2 |
| Any CM | 3,705 | 49.1 | 206,240 | 45.7 |

*Supplementary Table 4. Counts and frequency for each of the conditions of interest in individuals with and without a ND-CNV.*

| | OR (CI) | p-value |
|---|---|---|
| Anxiety | 1.25 (1.16-1.34) | $9.16x10^{-10}$ |
| Depression | 1.24 (1.16-1.31) | $5.11x10^{-12}$ |
| Somatoform disorder | 1.13 (0.96-1.32) | 0.144 |
| Any internalising condition | 1.22 (1.15 -1.28) | $4.50x10^{-12}$ |
| Hypertension | 1.16 (1.10-1.22) | $1.20x10^{-8}$ |
| Obesity | 1.27 (1.18-1.36) | $1.04x10^{-11}$ |
| Chronic kidney disease | 1.10 (1.22-1.46) | $1.69x10^{-3}$ |
| Type II diabetes | 1.37 (1.27-1.48) | $2.43x10^{-17}$ |
| Dyslipidaemia | 1.33 (1.03-1.16) | $1.45x10^{-10}$ |
| Any cardiometabolic condition | 1.19 (1.13-1.24) | $5.37x10^{-12}$ |
| Any internalising and hypertension | 1.21 (1.13-1.31) | $5.55x10^{-7}$ |
| Any internalising and dyslipidemia | 1.27 (1.16-1.38) | $1.34x10^{-7}$ |
| Any internalising and obesity | 1.48 (1.33-.164) | $1.51x10^{-13}$ |
| Any internalising T2D | 1.57 (1.38-1.77) | $1.66x10^{-6}$ |
| Any internalising and CKD | 1.44 (1.24-1.66) | $1.09x10^{-6}$ |
| Any ICM-MM | 1.25 (1.17-1.34) | $3.14x10^{-11}$ |

*Supplementary Table 5. Association of individual conditions and ICM-MM phenotypes with the presence of a ND-CNV.*

| | OR (CI) | p-value |
|---|---|---|
| Any ICM-MM | 1.18 (1.10-1.26) | $1.32x10^{-6}$ |
| Any internalising and hypertension | 1.14 (1.05-1.23) | $9.56x10^{-4}$ |
| Any internalising and dyslipidemia | 1.20 (1.10-1.32) | $3.89x10^{-5}$ |
| Any internalising T2D | 1.41 (1.24-1.60) | $7.11x10^{-8}$ |
| Any internalising and CKD | 1.35 (1.17-1.56) | $6.53x10^{-5}$ |

*Supplementary Table 6. Association of ICM-MM phenotypes with the presence of a ND-CNV adjusting for BMI. Obesity was not included as an outcome in this analysis.*

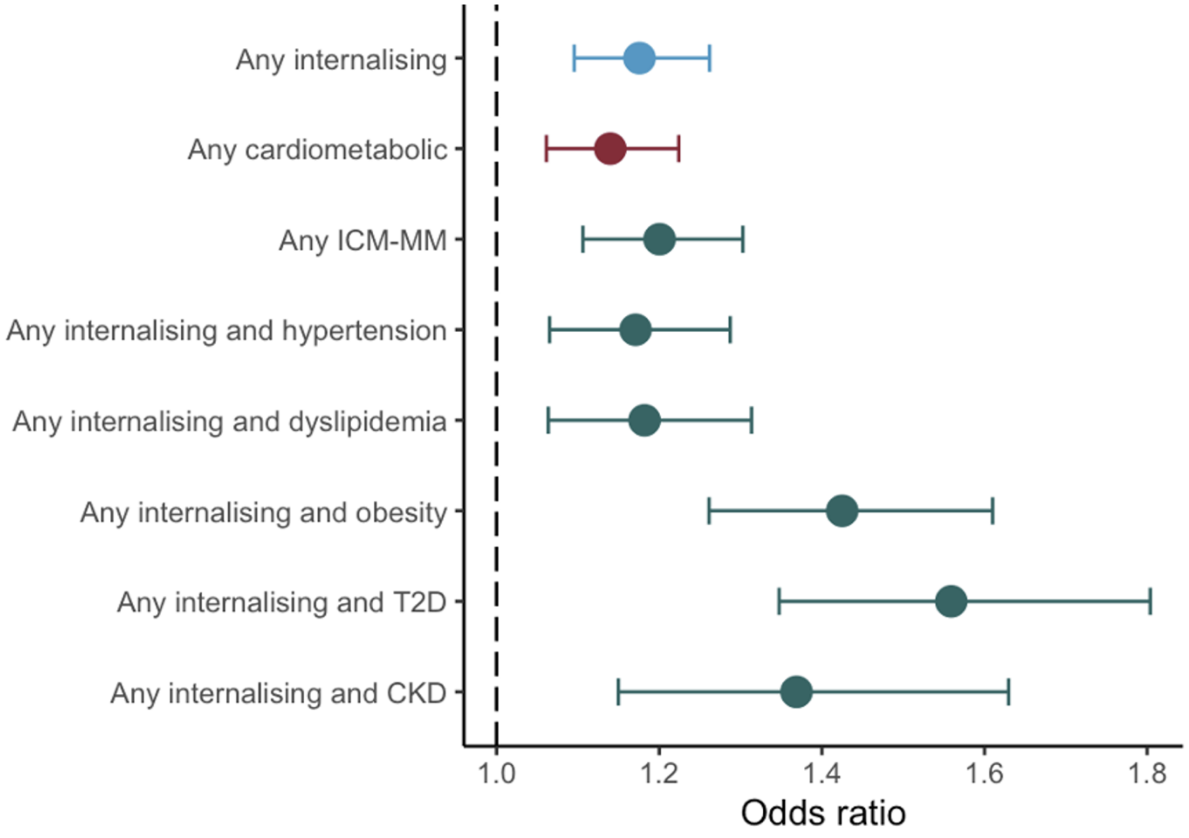


*Supplementary Figure 1. Association of ND-CNVs with multimorbidity for individuals with both primary care and HES data (N= 229,951).*

| | OR (CI) | p-value |
|---|---|---|
| Any ICM-MM | 1.20 (1.11-1.30) | $1.21 \times 10^{-5}$ |
| Any internalising and hypertension | 1.17 (1.07-1.29) | $1.09 \times 10^{-3}$ |
| Any internalising and dyslipidemia | 1.18 (1.06-1.31) | $1.92 \times 10^{-3}$ |
| Any internalising and obesity | 1.48 (1.26-1.61) | $1.30 \times 10^{-8}$ |
| Any internalising T2D | 1.37 (1.35-1.80) | $2.42 \times 10^{-9}$ |
| Any internalising and CKD | 1.56 (1.15-1.63) | $4.20 \times 10^{-4}$ |

*Supplementary Table 7. Association of ICM-MM phenotypes with the presence of a ND-CNV for individuals with both primary care and HES data (N= 229,951).*

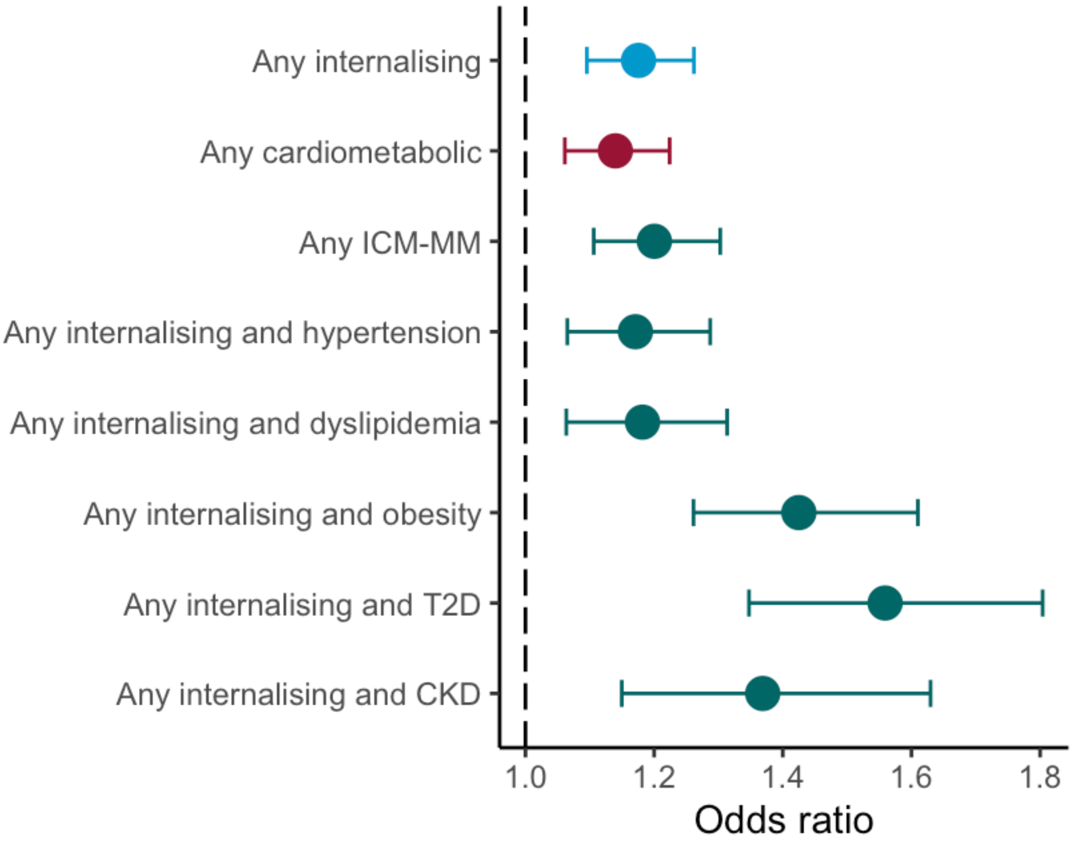


*Supplementary Figure 2. Association of ND-CNVs with multimorbidity excluding individuals with 16p11.2 proximal and distal deletions (N= 185).*

| | OR (CI) | p-value |
|---|---|---|
| Any ICM-MM | 1.20 (1.11-1.30) | $2.08x10^{-9}$ |
| Any internalising and hypertension | 1.17(1.07-1.29 | $7.64x10^{-6}$ |
| Any internalising and dyslipidemia | 1.18 (1.06-1.31) | $3.84x10^{-6}$ |
| Any internalising and obesity | 1.42 (1.26-1.61) | $8.95x10^{-10}$ |
| Any internalising T2D | 1.56 (1.35-1.80) | $7.06x10^{-10}$ |
| Any internalising and CKD | 1.37 (1.15-1.63) | $4.15x10^{-6}$ |

*Supplementary Table 8. Association of ICM-MM phenotypes with the presence of a ND-CNV excluding individuals with 16p11.2 proximal and distal deletions (N= 185).*

| | **Male sex** | **Female sex** | **Difference** |
|---|---|---|---|
| | **OR (CI)** | **OR (CI)** | **p-value** |
| Any internalising | 1.20 (1.10-1.31) | 1.25 (1.16-1.34) | 0.512 |
| Any cardiometabolic | 1.09 (1.02-1.17) | 1.25 (1.17-1.34) | 0.004 |
| Any ICM-MM | 1.19 (1.08-1.32) | 1.31 (1.20-1.43) | 0.211 |
| Any internalising and hypertension | 1.12 (0.99-1.26) | 1.31 (1.18-1.45) | 0.063 |
| Any internalising and dyslipidemia | 1.19 (1.05-1.36) | 1.34 (1.19-1.51) | 0.214 |
| Any internalising and obesity | 1.39 (1.17-1.66) | 1.55 (1.36-1.76) | 0.328 |
| Any internalising T2D | 1.35 (13.1-1.61) | 1.78 (1.51-2.10) | 0.026 |
| Any internalising and CKD | 1.61 (1.30-1.99) | 1.32 (1.08-1.62) | 0.181 |

*Supplementary Table 9. Association of ICM-MM phenotypes with ND-CNV for male and female sex.*

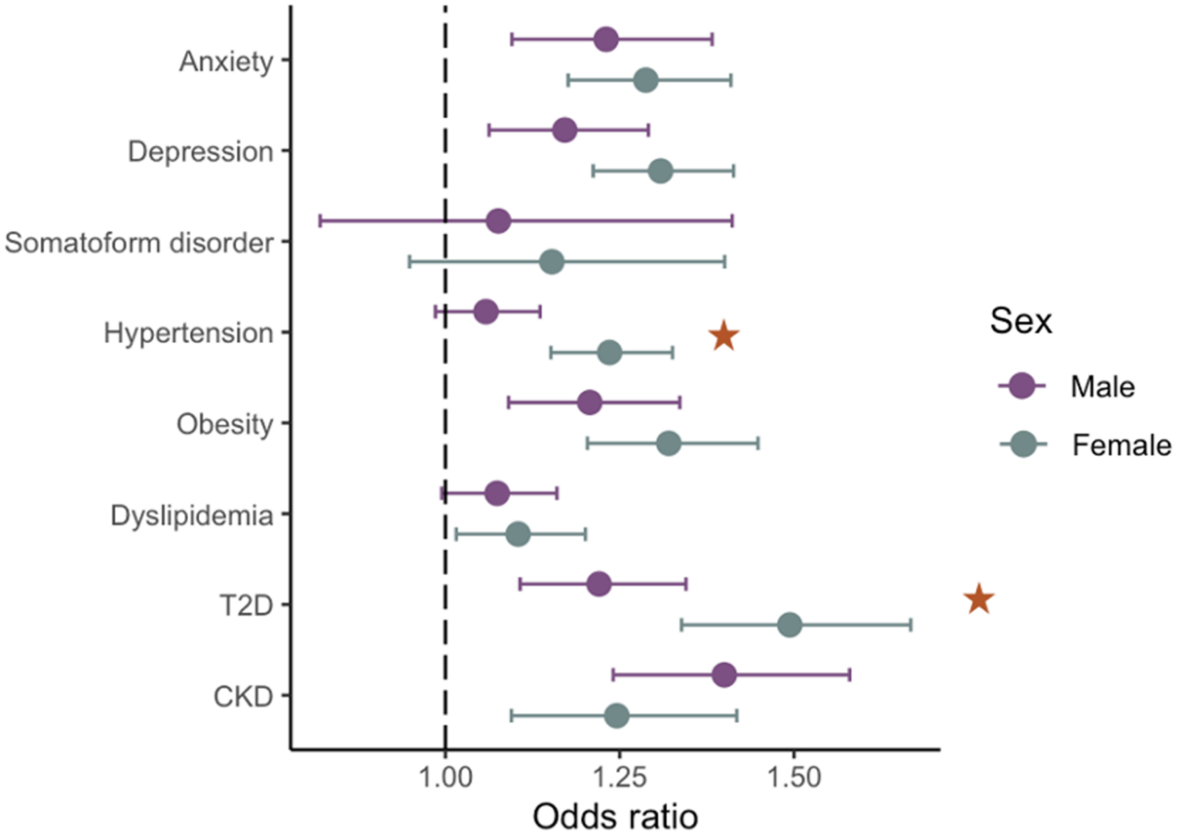


*Supplementary Figure 3. Association of ND-CNVs with internalising and cardiometabolic conditions for male and female sex. Stars indicate a significant interaction of presence of ND-CNV and sex.*

| | **Duplications** | **Deletions** | **Difference** |
|---|---|---|---|
| | **OR (CI)** | **OR (CI)** | **p-value** |
| Any internalising | 1.21 (1.13-1.29) | 1.26 (1.16-1.38) | 0.834 |
| Any cardiometabolic | 1.17 (1.10-1.24) | 1.17 (1.08-1.26) | 0.980 |
| Any ICM-MM | 1.21 (1.11-1.32) | 1.32 (1.19-1.47) | 0.430 |
| Any internalising and hypertension | 1.16 (1.05-1.28) | 1.31 (1.17-1.48) | 0.281 |
| Any internalising and dyslipidemia | 1.31 (1.18-1.47) | 1.20 (1.04-1.38) | 0.596 |
| Any internalising and obesity | 1.27 (1.10-1.46) | 1.85 (1.59-2.15) | 0.002 |
| Any internalising T2D | 1.45 (1.24-1.70) | 1.72 (1.44-2.07) | 0.267 |
| Any internalising and CKD | 1.34 (1.11-1.62) | 1.61 (1.29-2.02) | 0.439 |

*Supplementary Table 10. Association of ICM-MM phenotypes with duplications and deletions.*

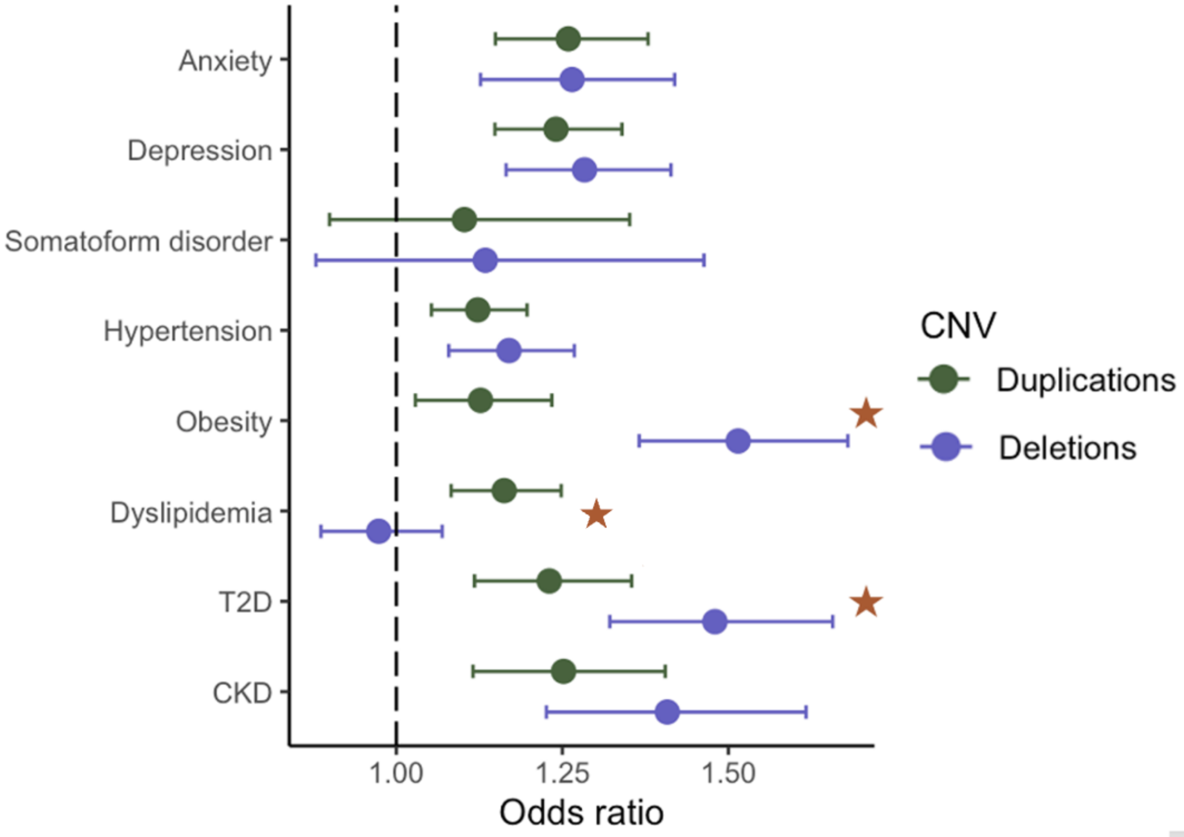


*Supplementary Figure 4. Association of duplications and deletions with internalising and cardiometabolic conditions. Stars indicate a suggested difference between deletions and duplications (p-value < 0.05).*

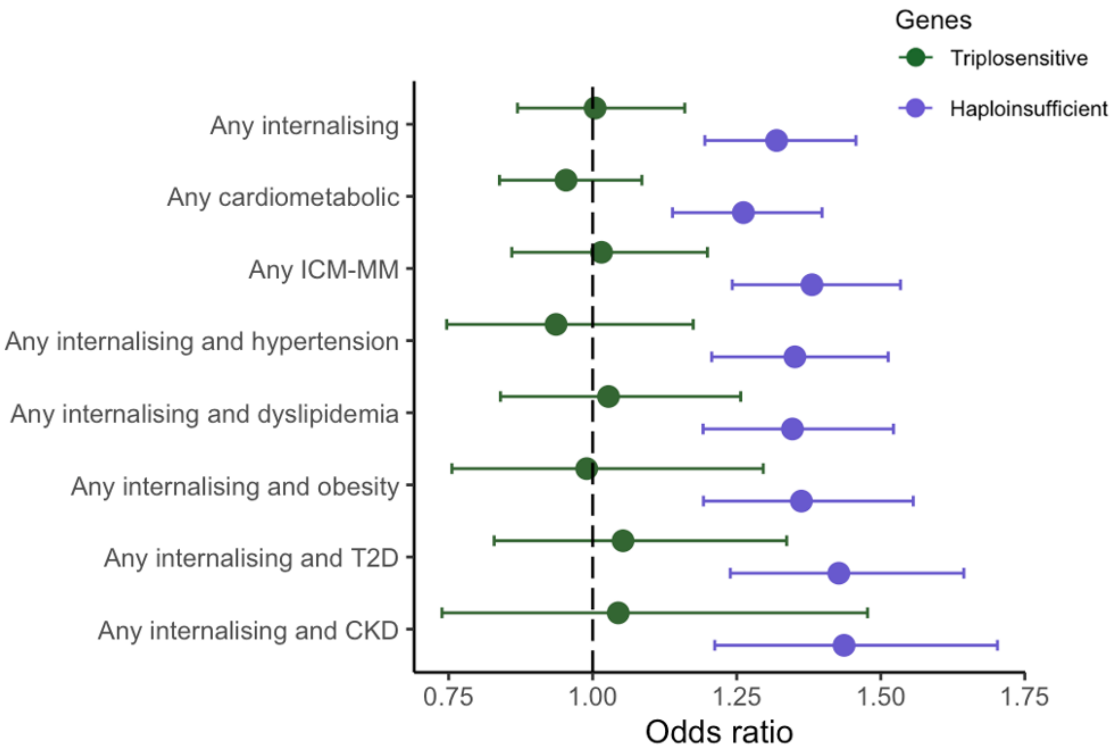


*Supplementary Figure 5. Association of triplosensitive genes in duplications and haploinsufficient genes in deletions with ICM-MM. ORs represent an increase in risk by an increase of one gene*

| | **Deletions** | | **Duplications** | |
|---|---|---|---|---|
| | **Haploinsufficient genes** | **Total genes** | **Triplosensitive genes** | **Total genes** |
| | **OR (CI)** | **OR (CI)** | **OR (CI)** | **OR (CI)** |
| Any internalising | 1.31 (1.19-1.46) * | 1.01 (1.00-1.01) * | 1.01 (0.86-1.15) | 1.01 (1.00-1.01) * |
| Any cardiometabolic | 1.26 (1.14-1.40) * | 1.01 (1.00-1.01) * | 0.95 (0.84-1.08) | 1.01 (1.00-1.01) * |
| Any ICM-MM | 1.38 (1.24-1.53) * | 1.01 (1.00-1.01) * | 1.01 (0.86 -1.20) | 1.01 (1.00-1.01) * |
| Any internalising and hypertension | 1.35 (1.21-1.51) * | 1 (1.00-1.01) | 0.94 (0.75-1.17) | 1. 01 (1.00-1.01) |
| Any internalising and dyslipidemia | 1.35 (1.19-1.52) * | 1.01 (1.00-1.01) * | 1.03 (0.84-1.25) | 1.01 (1.00-1.01) * |
| Any internalising and obesity | 1.36 (1.10-1.55) * | 1.01 (1.01-1.03) * | 0.99 (0.76-1.30) | 1.01 (1.00-1.01) * |
| Any internalising and T2D | 1.43 (1.24.1.64) * | 1.01 (1.01-1.02) * | 1.05 (0.83-1.34) | 1.01 (1.00-1.01) * |
| Any internalising and CKD | 1.44 (1.21-1.70) * | 1.01 (1.00-1.01) | 1.04 (0.74-1.48) | 1.01 (1.00-1.01) |

*Supplementary Table 11. Association of haploinsufficient and total genes in deletions and triplosensitive and total genes in duplications with ICM-MM. ORs represent odds by an increase of one gene. Asterisks indicate statistical significance after multiple testing.*

| PRS | MDD | | Anxiety | | T2D | | BMI | | SBP | | LDL | | CKD | |
|---|---|---|---|---|---|---|---|---|---|---|---|---|---|---|
| | OR (CI) | p-value | OR (CI) | p-value | OR (CI) | p-value | OR (CI) | p-value | OR (CI) | p-value | OR (CI) | p-value | OR (CI) | p-value |
| **Any ICM-MM** | 1.01 (0.94, 1.08) | 0.857 | 1.03 (0.96,1.10) | 0.438 | 1.00 (0.93,1.08) | 0.983 | 0.98 (0.91,1.06) | 0.668 | 0.96 (0.89,1.03) | 0.269 | 1.03 (0.96,1.11) | 0.351 | 1.02 (0.95,1.09) | 0.606 |
| **Any internalising and hypertension** | 0.96 (0.8, 81.04) | 0.323 | 1.01 (0.93,1.09) | 0.842 | 1.02 (0.94,1.11) | 0.616 | 1.01 (0.92,1.10) | 0.902 | 0.98 (0.90,1.07) | 0.678 | 1.06 (0.97,1.14) | 0.183 | 1.01 (0.93,1.10) | 0.775 |
| **Any internalising and dyslipidemia** | 1.04 (0.94, 1.14) | 0.468 | 1.07 (0.97,1.17) | 0.176 | 1.01 (0.92,1.12) | 0.795 | 0.97 (0.87,1.07) | 0.500 | 1.02 (0.93,1.12) | 0.651 | 1.05 (0.95,1.15) | 0.332 | 1.03 (0.94,1.13) | 0.535 |
| **Any internalising and obesity** | 1.10 (0.98, 1.23) | 0.094 | 1.15 (1.04,1.28) | 0.010 | 0.97 (0.87,1.09) | 0.642 | 0.98 (0.87,1.10) | 0.738 | 1.02 (0.92,1.15) | 0.667 | 1.02 (0.91,1.13) | 0.748 | 1.07 (0.96,1.19) | 0.237 |
| **Any internalising and T2D** | 0.93 (0.82, 1.07) | 0.317 | 1.01 (0.89,1.14) | 0.884 | 1.01 (0.88,1.16) | 0.907 | 0.98 (0.85,1.13) | 0.775 | 1.09 (0.96,1.25) | 0.177 | 1.05 (0.93,1.20) | 0.428 | 0.95 (0.84,1.08) | 0.413 |
| **Any internalising and CKD** | 1.06 (0.90, 1.24) | 0.486 | 1.18 (1.01,1.37) | 0.037 | 0.88 (0.75,1.04) | 0.143 | 0.94 (0.79,1.11) | 0.470 | 1.05 (0.90,1.23) | 0.549 | 1.18 (1.01,1.37) | 0.039 | 1.01 (0.87,1.18) | 0.893 |

*Supplementary Table 12. Interaction of ND-CNVs and PRSs.*

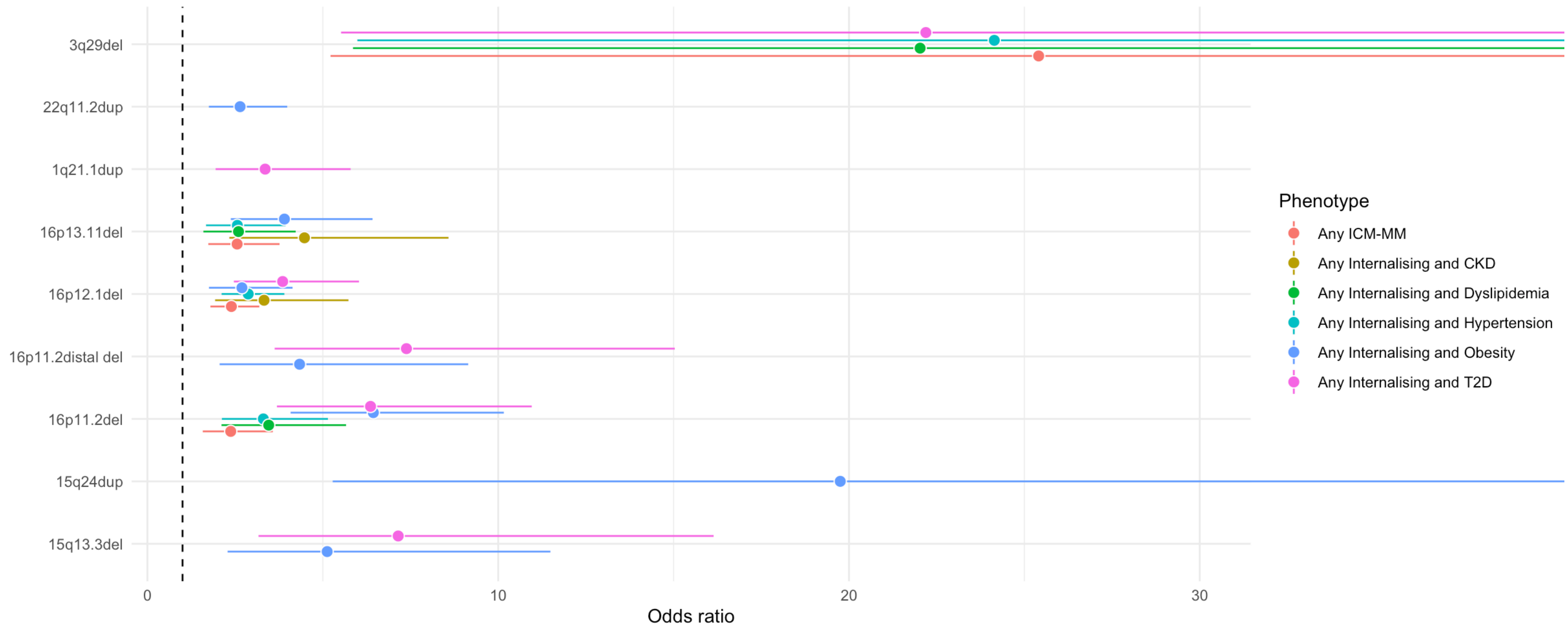


*Supplementary Figure 6. Association of individual ND-CNVs and multimorbidity. Only significant associations after Bonferroni correction for multiple testing are included in the figure.*

| | Any internalising and hypertension | | | Any internalising and dyslipidemia | | | Any internalising and obesity | | | Any internalising and T2D | | | Any internalising and CKD | | | Any ICM-MM | | |
|---|---|---|---|---|---|---|---|---|---|---|---|---|---|---|---|---|---|---|
| CNV | OR | CI | p-value | OR | CI | p-value | OR | CI | p-value | OR | CI | p-value | OR | CI | p-value | OR | CI | p-value |
| TAR del | 1.22 | 0.59,2.54 | 0.593 | 1.07 | 0.43,2.66 | 0.881 | 0.35 | 0.05,2.51 | 0.295 | 1.09 | 0.27,4.46 | 0.9 | 2.15 | 0.68,6.87 | 0.195 | 0.97 | 0.48,1.96 | 0.941 |
| TAR dup | 1.1 | 0.79,1.51 | 0.574 | 1.3 | 0.91,1.84 | 0.145 | 1.55 | 1.03,2.34 | 0.037 | 1.8 | 1.14,2.86 | 0.012 | 1.27 | 0.68,2.38 | 0.457 | 1.39 | 1.08,1.8 | 0.012 |
| 1q21.1 del | 0.83 | 0.41,1.71 | 0.617 | 0.74 | 0.3,1.83 | 0.52 | 1.71 | 0.8,3.68 | 0.168 | 1.13 | 0.36,3.55 | 0.839 | 1.04 | 0.26,4.24 | 0.954 | 0.92 | 0.52,1.65 | 0.786 |
| 1q21.1 dup | 1.93 | 1.29,2.89 | 0.001 | 2.26 | 1.46,3.51 | 2.66E-04 | 1.51 | 0.8,2.86 | 0.206 | 3.36 | 1.94,5.79 | 1.40E-05 | 1.87 | 0.83,4.24 | 0.133 | 1.65 | 1.14,2.38 | 0.007 |
| NRXN1 del | 1.2 | 0.72,1.98 | 0.487 | 0.77 | 0.38,1.57 | 0.47 | 2.08 | 1.16,3.75 | 0.014 | 1.94 | 0.95,3.96 | 0.069 | 0.64 | 0.16,2.6 | 0.536 | 1.33 | 0.87,2.02 | 0.188 |
| 2q11.2 del | 0.83 | 0.2,3.47 | 0.795 | 0.59 | 0.08,4.35 | 0.606 | 3.86 | 1.35,11.01 | 0.012 | 0 | 0,2.34E+43 | 0.875 | 0 | 0,4.83E+42 | 0.881 | 1.05 | 0.37,3 | 0.933 |
| 2q13 del | 1.74 | 0.79,3.87 | 0.171 | 1 | 0.31,3.23 | 0.993 | 2.2 | 0.79,6.09 | 0.13 | 0 | 0,7.69E+32 | 0.839 | 1.19 | 0.16,8.7 | 0.861 | 1.27 | 0.6,2.7 | 0.539 |
| 2q13 dup | 0.66 | 0.24,1.82 | 0.425 | 0.98 | 0.36,2.69 | 0.965 | 0.39 | 0.05,2.8 | 0.349 | 1.84 | 0.58,5.86 | 0.302 | 0.82 | 0.11,5.96 | 0.848 | 0.84 | 0.38,1.84 | 0.662 |
| 3q29 del | 24.14 | 5.98,97.42 | 7.71E-06 | 22.03 | 5.86,82.82 | 4.73E-06 | 3.64 | 0.45,29.32 | 0.224 | 22.19 | 5.52,89.21 | 1.26E-05 | 19.24 | 3.88,95.44 | 2.96E-04 | 25.41 | 5.22,123.68 | 6.17E-05 |
| WBS dup | 0.79 | 0.1,5.99 | 0.817 | 2.4 | 0.54,10.66 | 0.251 | 1.86 | 0.24,14.13 | 0.549 | 2.41 | 0.31,18.66 | 0.399 | 4.41 | 0.57,34.04 | 0.155 | 1 | 0.22,4.48 | 1 |
| 8p23.1 dup | 1.33 | 0.16,10.86 | 0.789 | 1.8 | 0.22,14.78 | 0.584 | 4.05 | 0.49,33.25 | 0.192 | 9.54 | 1.84,49.56 | 0.007 | 0 | 0,5.85E+32 | 0.872 | 4.49 | 1.07,18.9 | 0.041 |
| 15q11.2 del | 0.97 | 0.82,1.15 | 0.737 | 0.94 | 0.77,1.16 | 0.557 | 1.23 | 0.97,1.56 | 0.08 | 1.05 | 0.78,1.43 | 0.734 | 1.24 | 0.9,1.71 | 0.182 | 1 | 0.87,1.16 | 0.952 |
| 15q11.2 dup | 1.15 | 1,1.33 | 0.047 | 1.34 | 1.14,1.56 | 2.67E-04 | 1.22 | 1,1.5 | 0.054 | 1.36 | 1.07,1.71 | 0.01 | 1.21 | 0.91,1.61 | 0.2 | 1.15 | 1.02,1.3 | 0.024 |
| PWS dup | 0.81 | 0.11,6.11 | 0.837 | 1.22 | 0.16,9.22 | 0.848 | 0 | 0,4.69E+34 | 0.854 | 0 | 0,8.42E+34 | 0.866 | 0 | 0,1.378E+34 | 0.873 | 0.44 | 0.06,3.31 | 0.424 |
| 15q13.3 del | 3.06 | 1.51,6.19 | 0.002 | 3.86 | 1.85,8.05 | 3.30E-04 | 5.12 | 2.28,11.49 | 7.35E-05 | 7.15 | 3.17,16.14 | 2.21E-06 | 1.26 | 0.17,9.17 | 0.823 | 2.16 | 1.11,4.21 | 0.023 |
| 15q24 dup | 6.28 | 1.53,25.78 | 0.011 | 5.38 | 1.09,26.55 | 0.039 | 19.75 | 5.28,73.87 | 9.31E-06 | 6.33 | 0.79,50.99 | 0.083 | 8.28 | 0.98,70 | 0.052 | 6.17 | 1.63,23.37 | 0.007 |
| 16p13.11 del | 2.56 | 1.67,3.94 | 1.75E-05 | 2.6 | 1.59,4.23 | 1.25E-04 | 3.9 | 2.38,6.42 | 7.68E-08 | 2.65 | 1.29,5.41 | 0.008 | 4.48 | 2.33,8.58 | 6.45E-06 | 2.56 | 1.74,3.77 | 2.06E-06 |
| 16p13.11 dup | 1.18 | 0.94,1.47 | 0.157 | 1.32 | 1.03,1.7 | 0.029 | 1.18 | 0.84,1.65 | 0.336 | 1.71 | 1.22,2.41 | 0.002 | 1.63 | 1.09,2.43 | 0.016 | 1.09 | 0.89,1.33 | 0.392 |
| 16p12.1 del | 2.87 | 2.11,3.9 | 1.63E-11 | 1.86 | 1.24,2.8 | 0.003 | 2.69 | 1.75,4.14 | 6.14E-06 | 3.85 | 2.46,6.03 | 3.54E-09 | 3.32 | 1.93,5.73 | 1.54E-05 | 2.39 | 1.8,3.19 | 2.66E-09 |
| 16p11.2 distal del | 1.33 | 0.57,3.1 | 0.504 | 2.7 | 1.28,5.69 | 0.009 | 4.34 | 2.06,9.15 | 1.16E-04 | 7.38 | 3.63,15.03 | 3.58E-08 | 2.39 | 0.58,9.86 | 0.227 | 1.81 | 0.95,3.42 | 0.069 |
| 16p11.2 distal dup | 2.06 | 1.29,3.29 | 0.002 | 1.92 | 1.1,3.34 | 0.022 | 0.8 | 0.29,2.16 | 0.654 | 0.64 | 0.16,2.57 | 0.527 | 1.35 | 0.43,4.27 | 0.605 | 1.77 | 1.16,2.71 | 0.008 |
| 16p11.2 del | 3.3 | 2.12,5.15 | 1.24E-07 | 3.46 | 2.11,5.66 | 8.42E-07 | 6.44 | 4.08,10.16 | 1.20E-15 | 6.36 | 3.69,10.96 | 2.63E-11 | 3.21 | 1.3,7.93 | 0.011 | 2.37 | 1.58,3.57 | 3.46E-05 |
| 16p11.2 dup | 0.76 | 0.38,1.49 | 0.417 | 0.84 | 0.39,1.81 | 0.665 | 0.61 | 0.19,1.91 | 0.396 | 1.25 | 0.46,3.37 | 0.665 | 2.09 | 0.85,5.13 | 0.108 | 1.04 | 0.63,1.71 | 0.89 |
| 17p13.3 YWHAE del | 0.98 | 0.23,4.16 | 0.979 | 0.69 | 0.09,5.13 | 0.72 | 4.96 | 1.71,14.45 | 0.003 | 3.69 | 0.87,15.65 | 0.076 | 2.7 | 0.36,20.11 | 0.333 | 1.41 | 0.48,4.1 | 0.533 |
| 17p13.3 YWHA Edup | 3.68 | 0.73,18.59 | 0.115 | 2.25 | 0.27,18.56 | 0.452 | 4.7 | 0.57,38.44 | 0.149 | 5.73 | 0.7,47.05 | 0.104 | 7.64 | 0.9,64.72 | 0.062 | 1.16 | 0.14,9.84 | 0.892 |
| Potocki Lupski | 2.25 | 0.26,19.47 | 0.461 | 0 | 0,0 | 0.818 | 0 | 0,1.54E+38 | 0.88 | 0 | 0,2.59E+38 | 0.888 | 0 | 0,1.47E+38 | 0.893 | 1.47 | 0.17,12.77 | 0.727 |
| 17q11.2 del NF1 | 0 | 0,0 | 0.75 | 0 | 0,2.97E+28 | 0.833 | 3.34 | 0.42,26.52 | 0.255 | 0 | 0,1.01E+29 | 0.854 | 0 | 0,6.32E+48 | 0.901 | 0.88 | 0.11,6.98 | 0.902 |
| 17q12 del | 3.85 | 0.79,18.74 | 0.095 | 5.46 | 1.12,26.71 | 0.036 | 0 | 0,4.04E+30 | 0.857 | 12.9 | 2.64,63.12 | 0.002 | 10.28 | 1.25,84.54 | 0.03 | 2.11 | 0.43,10.23 | 0.355 |
| 17q12 dup | 1.49 | 0.81,2.73 | 0.196 | 0.82 | 0.33,2.02 | 0.665 | 1.84 | 0.8,4.2 | 0.149 | 1.21 | 0.38,3.82 | 0.747 | 2.99 | 1.21,7.41 | 0.018 | 1.3 | 0.75,2.26 | 0.352 |
| 22q11.2 del | 3.74 | 0.79,17.73 | 0.097 | 0 | 0,5.75E+28 | 0.843 | 3.22 | 0.41,25.62 | 0.269 | 0 | 0,1.47E+29 | 0.861 | 0 | 0,2.05E+49 | 0.91 | 1.79 | 0.37,8.6 | 0.468 |
| 22q11.2 dup | 1.26 | 0.85,1.87 | 0.244 | 1.5 | 0.98,2.3 | 0.063 | 2.64 | 1.75,3.98 | 3.70E-06 | 1.75 | 0.96,3.2 | 0.069 | 0.87 | 0.32,2.35 | 0.79 | 1.23 | 0.88,1.71 | 0.22 |
| 22q11.2 distal del | 0 | 0,0 | 0.829 | 0 | 0,3.76E+24 | 0.839 | 0 | 0,5.47E+24 | 0.846 | 0 | 0,2.86E+42 | 0.901 | 0 | 0,8.65E+41 | 0.907 | 0 | 0,5.80E+23 | 0.808 |
| 22q11.2 distal dup | 1.77 | 0.39,7.93 | 0.458 | 1.19 | 0.15,9.12 | 0.868 | 0 | 0,8.99E+23 | 0.821 | 0 | 0,1.95E+41 | 0.881 | 0 | 0,1.04E+41 | 0.884 | 1.08 | 0.24,4.9 | 0.923 |
| | | | | | | | | | | | | | | | | | | 0.593 |

*Supplementary Table 13. Association of each of the ND-CNVs with ICM-MM.*
